# Robot-based assessment of HIV-related motor and cognitive impairment for neurorehabilitation

**DOI:** 10.1101/2020.10.30.20223172

**Authors:** Kevin D. Bui, Carol A. Wamsley, Frances S. Shofer, Dennis L. Kolson, Michelle J. Johnson

## Abstract

There is a pressing need for strategies to slow or treat the progression of functional decline in people living with HIV. This paper explores a novel rehabilitation robotics approach to measuring cognitive and motor impairment in adults living with HIV, including a subset with stroke. We conducted a cross-sectional study with 21 subjects exhibiting varying levels of cognitive and motor impairment. We developed three robot-based tasks trajectory tracking, N-back, and spatial span – to assess if metrics derived from these tasks were sensitive to differences in subjects with varying levels of executive function and upper limb motor impairments. We also examined if these metrics could estimate clinical cognitive and motor scores. The results showed that the average sequence length on the robot-based spatial span task was the most sensitive to differences between subjects’ cognitive and motor impairment levels. We observed strong correlations between robot-based measures and clinical cognitive and motor assessments relevant to the HIV population, such as the Color Trails 1 (rho = 0.83), Color Trails 2 (rho = 0.71), Digit Symbol – Coding (rho = 0.81), Montreal Cognitive Assessment – Executive Function subscore (rho = 0.70), and Box and Block Test (rho = 0.74). Importantly, our results highlight that gross motor impairment may be overlooked in the assessment of HIV-related disability. This study shows that rehabilitation robotics can be expanded to new populations beyond stroke, namely to people living with HIV and those with cognitive impairments.

## I. Introduction

Today, there are nearly 37 million persons living with human immunodeficiency virus (PLWHIV) worldwide [1]. As PLWHIV age due to the success of antiretroviral therapy (ART), the challenges have shifted to managing the chronic effects of living with HIV. Many of these challenges can be attributed to neurological complications caused by HIV-associated neurocognitive disorders (HAND), accelerated aging, drug abuse, and HIV-related comorbidities [2]. Together, the broad range of impairments experienced by PLWHIV has been shown to impact instrumental activities of daily living (IADLs), such as medication management, telephone communication, cooking, and financial management [3]. In one study, upwards of 80% of PLWHIV reported dealing with at least one impairment, activity limitation or disability, or social participation restriction [4]. These deficits are often tied to impairments in executive function, memory, and visuospatial domains [5]. PLWHIV also experience motor impairments in gait, coordination, upper limb fine motor skills, and strength, with 69% of PLWHIV in one study demonstrating at least one motor impairment [6]–[9]. As such, there is a pressing need for effective neurorehabilitation strategies to slow or treat the progression of functional decline in PLWHIV.

The gold standard for diagnosing neurocognitive impairment has been established by the Frascati criteria, an extensive neuropsychological battery that classifies HAND subtypes as asymptomatic neurocognitive impairment, mild neurocognitive disorder, or HIV-associated dementia [10]. However, the assessments used to diagnose HAND often test domains in isolation, which is not reflective of the dual involvement of cognitive and motor demands in most IADL tasks. Differences between HIV and non-HIV populations are also seen in more nuanced tasks. Kronemer et al. demonstrated that even when there was no motor impairment detected on clinical assessments, PLWHIV demonstrated upper limb motor impairment while multitasking compared to a non-HIV control group that were not related to HAND stage [11]. Assessments of multitasking have been shown to be more reflective of IADL performance in PLWHIV compared to standard clinical assessments [12]. These results that current clinical assessments and biomarkers of HIV do not necessarily correspond well to more subtle impairments in cognition and motor performance [11].

HIV-associated non-communicable diseases, such as cerebrovascular disease (CVD), are a secondary effect of HIV infection that can further exacerbate existing cognitive and motor impairments. HIV is an independent risk factor for CVD such as stroke [13]. With an incidence rate of 3.87 per 1000 years lived, CVDs occur at an average age of 48 years in the HIV population [13]. These numbers are 1.5 times higher and 22 years younger than the general U.S. population [14]. Augustyn et al. recently showed that stroke survivors with HIV experienced a decline in ADL functions one month after discharge compared to stroke survivors without HIV who continued to show improvement, highlighting how HIV can impact stroke recovery [15].

Efforts to develop neurorehabilitation strategies have been made in the stroke population, but there is a paucity of established solutions for PLWHIV despite evidence that rehabilitation can positively address HIV-related challenges in physical, social, and psychological well-being [16], [17]. The rehabilitation robotics field provides a potential solution to address these challenges [18]. Robot-assisted stroke therapy has been shown to be as effective as high-intensity physical therapy for chronic stroke patients [19]. Additionally, robotic systems allow for a variety of kinematic metrics to be observed that relate to clinical measures of motor impairment [20]–[24].

While the primary focus to date has been on motor impairment, recent studies have started to look at robot-based measures of cognitive impairment in stroke and traumatic brain injury populations [21], [22]. Both of these studies have demonstrated a relationship between robot-based metrics and overall cognitive scores. However, given that cognition is broadly defined, more work needs to be done to establish robot-based metrics relating to specific domains.

The strengths of a rehabilitation robotics-based approach include the ability to standardize assessments with a greater range of objective measures, collect a vast amount of data, and develop personalized neurorehabilitation strategies based on the patient’s presenting characteristics. Prior work has also shown the feasibility of deploying cost-effective rehabilitation robotics systems in lower-resource contexts [25]. Cost-effective rehabilitation robotics system can bridge healthcare gaps in countries with low-to-middle income economies that are dealing with large populations of patients with impairments and a shortage of rehabilitation professionals. This approach has the potential to positively impact PLWHIV by building upon the body of work that has been done in the stroke population.

This preliminary cross-sectional study aims to establish objective, robot-based measures of executive function and upper limb motor impairment in PLWHIV – including a subset with stroke – and assess the strength of the relationship between these robot-based and clinical assessment scores. This study tests three hypotheses to demonstrate the utility of a robotic approach in assessing impairments in PLWHIV. Given the heterogeneous nature of impairments, the first part of this study tests the hypothesis that robot-based metrics can differentiate subjects with and without moderate executive function or upper-limb motor impairments (H1). The second hypothesis measures the relationship between robot-based metrics and clinical assessments used in PLWHIV by testing the hypothesis that robot-based metrics are good predictors of clinical cognitive assessment scores (H2) as well as clinical motor assessment scores (H3). This work lays the foundation for development of novel neurorehabilitation strategies for PLWHIV.

## II. Methods

### A. Subject population and procedure

Individuals over the age of 18 years old were recruited from the community through flyers posted at local HIV clinics and organizations. Inclusion criteria for the HIV group consisted of documented HIV status that was ART-treated and virally-suppressed, the ability to ambulate, the ability to comprehend study procedures, and the ability to provide written informed consent. Individuals with neuropathy (i.e. distal symmetric polyneuropathy) were excluded.

Subjects were included in the HIV-stroke subgroup if they met the inclusion criteria for the HIV group and were at least three months removed from a stroke event. HIV-stroke subjects with severe aphasia, visual neglect, or basal ganglia stroke were excluded. Subjects were excluded if they were more than mildly depressed as assessed by the Beck’s Depression Inventory – Fast Screen (score ≥ 4) [26]. Subjects were compensated for time and travel. This protocol was approved by the Internal Review Board of the University of Pennsylvania (Protocol no. 823511).

Subjects underwent a preliminary phone screen to screen for study eligibility. They were then sent a copy of the informed consent to review prior to coming in for their scheduled in-person appointment. After written informed consent were obtained in-person, cognitive and motor assessments were performed. Participants then completed three robot-based tasks in a randomized order with the dominant and non-dominant upper-extremity limb.

### B. Cognitive assessments

The cognitive assessments consisted of the Color Trails, Digit Symbol–Coding (WAIS-III ®), Montreal Cognitive Assessment (MoCA), and International HIV Dementia Scale (IHDS) [27]–[30]. These tests have all been administered in PLWHIV previously to measure neurocognitive impairment [8], [30]–[33]. These tests were chosen to reflect the domains that are commonly affected by HIV.

#### 1) Color Trails

The Color Trails is a set of two cognitive pencil and paper tests based on the Trail Making Test but does not require knowledge of the alphabet, thus reducing potential bias [27]. Color Trails 1 tests for sustained visual attention and simple sequencing, while Color Trails 2 assesses frontal systems such as selective attention, mental flexibility, visual spatial skills, and motor speed. Performance was measured by the time to complete the task, with a higher time indicating worse performance. These scores were normalized by age, gender, and education [27].

#### 2) Digit Symbol – Coding (WAIS-III ®)

The Digit Symbol–Coding (WAIS-III ®) test is another neuropsychological test assessing processing speed [28]. Subjects use a number-symbol key to copy symbols under a sequence of numbers. Performance was measured by the number of symbols coded in the span of two minutes, with a higher number of symbols copied in the time span representing better performance. Scores were normalized by age, gender, and education.

#### 3) Montreal Cognitive Assessment (MoCA)

The MoCA is a screening tool to detect impairment in a number of cognitive domains – visuospatial/executive, naming, memory, attention, language, abstraction, delayed recall, and orientation – and reflects the degree of cognitive impairment in a subject [29]. A score above 25 out of 30 generally indicates normal cognitive function, while a score below 19 indicates likely moderate cognitive impairment.

An executive function subscore (MoCA-EF) was calculated to serve as a proxy in place of a more extensive neuropsychological assessment of executive function, based on work by others demonstrating good convergent validity between this subscore and standardized neuropsychological tests of executive function [34]. This subscore, scored out of five points, was calculated from summing the scores from the backward digit span, trail making, word similarities, and ‘F’-word list generation tasks [34]. Lam et al. demonstrated that a cutoff score of 4 had a sesitivity of 0.79 to executive function impairment [34].

#### 4) International HIV Dementia Scale (IHDS)

The IHDS is a screening test for cognitive impairment designed to screen for HAND, with a score below 10 out of 12 indicating potential cognitive impairment [30]. It was developed as a culturally appropriate adaptation of the HIV Dementia Scale. However, the IHDS has not been validated in the stroke population.

### C. Motor assessments

The motor assessments tested gross motor function, fine motor function, and strength. They consisted of the Box and Blocks Test (BBT), Grooved Pegboard (GP), and grip strength.

#### 1) Box and Blocks (BBT)

The BBT is a test of gross motor function measuring how many blocks subjects are able to transfer across a partition in one minute, with a higher number of transferred blocks indicating better motor function [35]. Scores were normalized by age, gender, and limb. It is typically used to measure reach and grasp function in the stroke population.

#### 2) Grooved Pegboard (GP)

GP is a common assessment in PLWHIV. It tests fine motor function and dexterity and measures the amount of time a subject takes to insert all of the grooved pegs into matched holes on a board, with longer times to completion indicating worse fine motor function. Performance was measured by the time to complete the task [36]. GP data for subjects unable to complete the task were not included in the analysis (one subject).

#### 3) Grip Strength

Grip strength is measured with a dynamometer. Three trials were taken with each hand, with the average and standard deviation being recorded. Accelerated grip strength decline has been shown in a study of HIV-infected men, which may contribute to decreased life expectancy and lower quality of life with aging [37].

### D. Robot Assessment

#### 1) Rehabilitation Robot System

The rehabilitation robot used in this study, the Haptic TheraDrive, is a one degree-of-freedom robot for upper limb stroke rehabilitation (Fig. 1) [25]. The user operates the TheraDrive by manipulating a vertically-mounted crank handle equipped with force sensors and an optical encoder. For assessment purposes, it is run in a gravity-compensation mode, which uses force sensors as an input to a proportional-integral-derivative (PID) controller to calculate the necessary response by the motor to give the sensation that there is no resistance or assistance while the user manipulates the handle.

**Fig. 1.**
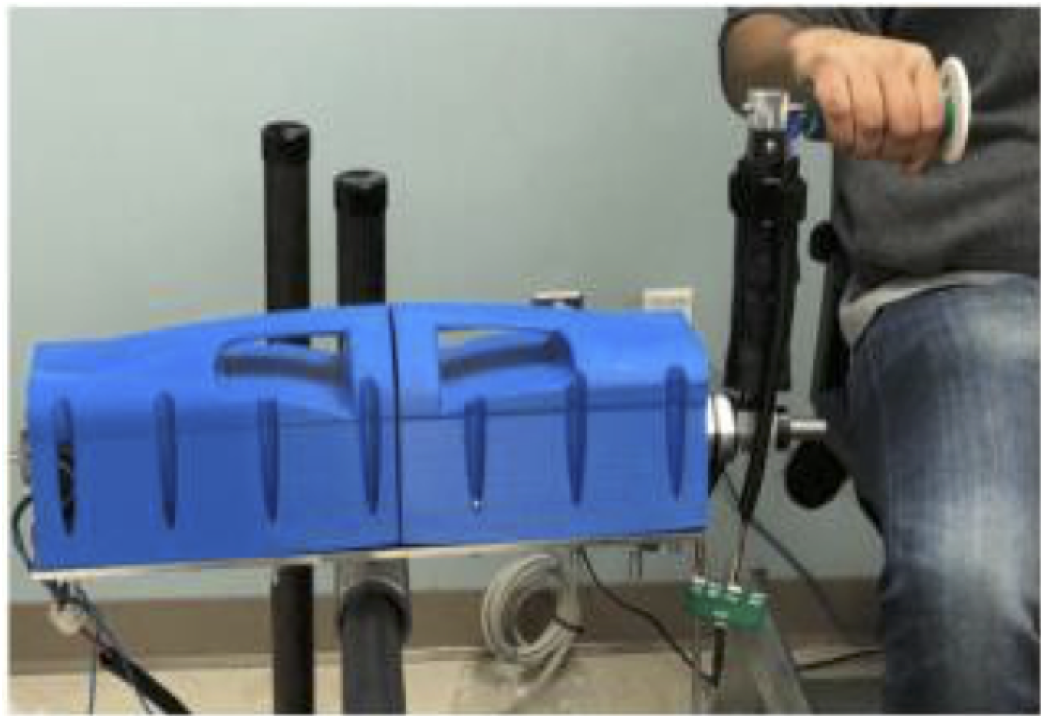
The Haptic Theradrive, a one degree-of-freedom rehabilitation robot system used in this study. Image used with permission from [38].

#### 2) Trajectory Tracking Motor Task

The trajectory tracking task is designed to assess upper limb motor performance. A single trial consists of the user moving the crank arm forward and backward to follow a a vertically scrolling sinusoidal path for 15 seconds. This task is repeated 15 times after one training trial. The outcome measures include performance error, movement smoothness, and the normalized distance traversed. Performance error was calculated as the root mean square error (RMSE) of the position relative to the displayed trajectory and normalized by the RMSE assuming no movement. A lower performance error indicated better tracking performance. Spectral arc length was used as the measure of smoothness, which has the benefit of being less sensitive to noise compared to other measures of smoothness [39]. More negative values of smoothness indicate less smooth movements. Normalized distance traversed was calculated from dividing the total angular distance that the subject traversed by the expected angular distance of the displayed trajectory path. A value closer to 1 reflects that the actual distance traversed matched the expected distance. A lower value could reflect moderate motor impairment, while a higher value could reflect inefficient movement.

#### 3) N-Back Cognitive Task

The N-back test is commonly used in the cognitive neuroscience field as a test of working memory and working memory capacity [40]. In this version, the subject is presented with a sequence of numerical digits (1-4) with three different conditions. For the 0-back condition, the easiest condition, the subject indicates when the current stimulus shown on the screen is the number ‘2.’ For the more cognitive involved 1-back and 2-back conditions, the subject indicates when the current stimulus matched the stimulus shown one stimulus or two stimuli prior, respectively. The subject indicates a match by pressing a button placed on the handle of the TheraDrive. The number would then flash green or red for a correct or incorrect response, respectively. Each subject performed the task with each limb, cycling through the 0-back, 1-back, and 2-back conditions four times for a total of 12 trials, all with different numerical sequences. The first set of trials is used as a training set and not included in the analysis. Ten responses are recorded per trial. Each subject was shown the same set of 12 sequences, with each sequence having a minimum of three button press responses. N-back performance was measured as the total number of correct responses divided by the total number of responses across the trials, resulting in a score ranging from 0 to 1, with 1 representing perfect performance.

#### 4) Spatial Span Cognitive-Motor Task

The Spatial Span is a test of visuospatial working memory based on the Corsi block-tapping task used in neuropsychological assessments [41]. While computerized versions of the Spatial Span exist [42], this version incorporates an added motor component to concurrently test for arm coordination, visuospatial ability, and working memory. A 3-by-3 grid of tiles is displayed to the user on a computer screen, and a sequence of tiles is shown one tile at a time. The user must repeat the order shown by operating the TheraDrive to select the tiles. If the user successfully repeats the sequence by selecting the correct tiles in order, the next displayed sequence increases in length by one to make the task more difficult. If the user is unsuccessful, the sequence decreases in length by one. The metrics of interest for the task include the normalized distance traversed, movement smoothness, mean sequence length across all the trials, and performance. Normalized distance traversed and movement smoothness were calculated the same way as in the trajectory tracking task. Mean sequence length is the average number of tiles displayed to the user across all the trials and reflects the capacity of the subject. Spatial span performance was measured as the total number correct tile matches divided by the total number of responses across the trials. Thus, spatial span performance is a score ranging from 0 to 1, with 1 representing perfect performance.

### E. Data processing

A one-sample Kolmogorov-Smirnov test for normal distribution was run on the raw continuous demographic, clinical, and robot metrics. Given that the data were not normally distributed, non-parametric Wilcoxon rank-sum tests were conducted to test for differences between HIV and HIV-stroke groups. To adjust for multiple comparisons, separate Bonferroni corrections were applied for the clinical (adjusted p = 0.004) and robot-based (adjusted p = 0.006) scores.

All robot metrics were Z-score normalized by the entire subject population in this study, resulting in a distribution with a mean of zero and standard deviation of one. This was done to ensure metrics were evenly weighted in the regression analysis.

### F. Functional subgroup comparison analysis

To investigate the first hypothesis that robot-based metrics can differentiate between subjects with and without moderate executive function impairments or upper-limb motor impairment, all study subjects were categorized by their motor and cognitive status based on clinical score cutoffs. The subject population demonstrated motor impairment on both the BBT and GP based on healthy population norms, but BBT was chosen to avoid excluding individual subjects who did not complete the GP. To categorize subjects by motor status, raw BBT scores were normalized by published gender, age, and limb side norms and converted into a Z-score. A BBT Z-score of -2 and below was used to indicate moderate motor impairment. To categorize subjects by cognitive status, a MoCA-EF score of 3.5 and below was used as a cutoff for likely moderate executive function impairment [34]. Subjects were then categorized into one of four functional subgroups based on the possible combinations of motor and cognitive status. Because this was done for both dominant and non-dominant limb motor status, subjects could be classified into two different functional subgroup classifications based on differing motor performance between dominant and non-dominant limbs.

For each robot-based metric, a two-way analysis of variance (ANOVA) was conducted where the factors were functional group and limb performance side. To adjust for all pairwise comparisons between functional groups, a Tukey-Kramer honest significance test was applied if the ANOVA was significant. An alpha level of 0.05 was used to establish the significance on all statistical tests.

### G. Multiple linear regression analysis

To investigate whether the robot-based metrics were significant predictors of clinical assessment scores, a multiple linear regression approach was used. Bosecker et al. previously used a backward multiple linear approach to identify a set of robot-based metrics reflective of various stroke outcome measures [23]. Rather than start with all of the robot-based metrics and remove terms, a forward stepwise approach was implemented. This consisted of individually testing each robot-based metric and subsequently adding it to the model only if it was a statistically significant predictor individually. Given the sample size of the subject population, the model was limited to two terms. In order to adjust for the number of predictors used in the model and compare between models with different numbers of predictors, the adjusted R^2^ is reported. A power analysis revealed that the linear regression models were powered to detect a minimum R^2^ of 0.40 with one predictor and 0.43 with two predictors (n = 21, power = 0.80, alpha = 0.05). A small, medium, and large effect size were defined as an R^2^ value of 0.01, 0.25, and 0.50, respectively. The non-parametric Spearman’s rho was also calculated to measure the correlation between predicted and actual clinical scores. All analysis was conducted in Matlab 2019A.

## III. Results

### A. Subject Population Breakdown

The descriptive statistics for demographic and clinical information for the subject groups (HIV, HIV-stroke, and combined) are presented in Table I. Twenty-one subjects in total – thirteen male and eight female – participated in the study. Six subjects had a history of stroke. The average age of the HIV and HIV-stroke groups were 56.2±5.4 years old and 54.2±8.1 years old, respectively, while the average age of the entire subject population was 55.5±6.3 years old. Fifteen subjects had 12 or more years of education. Fourteen subjects had MoCA-EF scores below 3.5 and sixteen subjects displayed moderate motor impairment in at least one limb based on MoCA-EF and BBT scores. There were no statistically significant differences – even at the unadjusted alpha level of 0.05 – between HIV and HIV-stroke groups on age or any of the clinical scores or between limbs on the clinical motor assessments.

**TABLE I.** Subject Demographics and Clinical Scores.

| Characteristics | HIV-only<br>mean±standard<br>deviation<br>(n = 15) | HIV-stroke<br>group<br>mean±standard<br>deviation<br>(n = 6) | Subject<br>population<br>mean±standard<br>deviation<br>(n = 21) |
| --- | --- | --- | --- |
| Age (years old) | 56.2 ± 5.4 | 54.2±8.1 | 55.5 ±6.3 |
| Gender<br>(Male/Female) | 10M/5F | 3M/3F | 13M/8F |
| $\geq 12$ years edu<br>(count) | 10 | 5 | 15 |
| Color Trails 1<br>(seconds) | 50.27±21.74 | 41.83±13.48 | 47.86±20.10 |
| Color Trails 2<br>(seconds) | 125.53±67.30 | 105.50±22.60 | 119.81±58.85 |
| Digit Symbol–<br>Coding Score | 45.07±12.48 | 49.67±6.16 | 46.38±11.24 |
| MoCA<br>(out of 30) | 21.47±4.43 | 23.83 ±2.19 | 22.14±4.06 |
| MoCA-EF<br>(score out of 5) | 2.87±1.31 | 2.83±0.69 | 2.86±1.17 |
| IHDS (out of<br>12) | 7.47±2.60 | 8.00±2.75 | 7.62±2.66 |
| Dominant<br>BBT (blocks) | 54.20±9.73 | 52.75±11.57 | 53.79±10.01 |
| Non-Dominant<br>BBT (blocks) | 54.40±9.54 | 47.83±18.62 | 52.52±12.63 |
| Dominant<br>GP (seconds) | 91.60±26.36 | 102.92±40.55 | 94.83±30.41 |
| Non-Dominant<br>GP (seconds) | 111.53±49.49 | 160.33±84.63 | 125.48±63.36 |
| Dominant Grip<br>Strength (kg) | 29.56±12.15 | 30.93±3.22 | 29.95±10.31 |
| Non-Dominant<br>Grip Strength<br>(kg) | 28.08±13.60 | 22.13±10.97 | 26.38±12.93 |

### B. Robot-Based Performance for Example Subjects

Performance data from two sample subjects (Subjects 12 and 18) on the trajectory tracking and spatial span tasks are presented, highlighting the wide variety of impairments seen in the subject population (Fig. 2). Subject 12 is a 56-year-old male HIV subject with moderate cognitive and moderate motor impairment, scoring a 13 on the MoCA and more than two standard deviations below Box and Block population norms on both the dominant and non-dominant limb. Subject 18 is a 49-year-old male HIV-stroke subject with low cognitive and low motor impairment, scoring a 25 on the MoCA and less than two standard deviations below BBT populations norms on both the dominant and non-dominant limb. Qualitatively, Subject 12 demonstrates poorer performance compared to Subject 18 (Fig. 2; left). This can be seen in comparing the average trajectory of each subject to the desired trajectory and the larger variance across the trials as seen in the shaded regions. On the robotic spatial span task, the distribution of sequence lengths across the task trials show a distinct difference between the two subjects (Fig. 2; right).

**Fig. 2.**
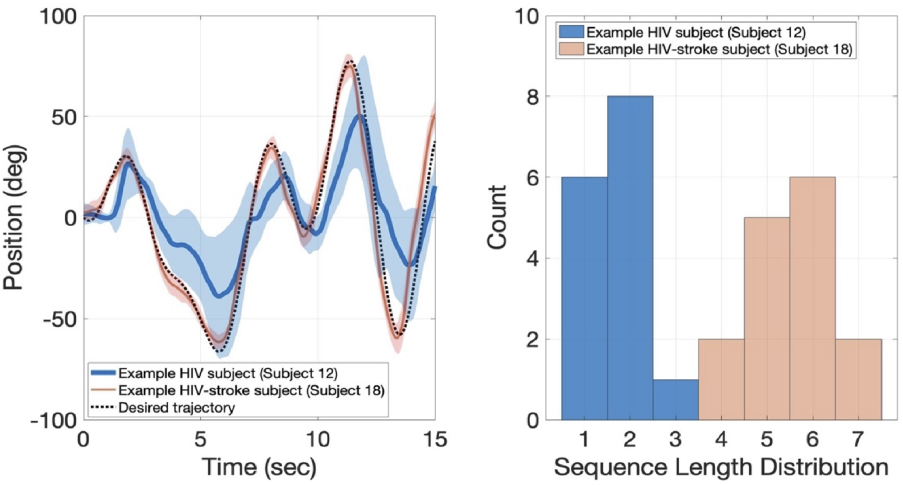
Left: The mean trajectory from the trajectory tracking task is shown for an example HIV (blue) and HIV-stroke (pink) subject. The expected trajectory is shown as a black dotted line. The shaded region represents the standard deviation across all the trials. Right: Histograms showing the distribution of sequence lengths on the spatial span task for the same HIV and HIV-stroke subject.

### C. Raw Robot Performance Metrics

Table II shows the mean and standard deviations for the raw robot-based metrics across the HIV-only group, HIV-stroke group, and the entire subject population. The scores for both the dominant and non-dominant limb are reported. There were no statistically significant differences – even at the unadjusted alpha level of 0.05 – in any robot metrics between dominant and non-dominant limbs or between HIV and HIV-stroke groups. However, some qualitative differences are notable. For example, while trajectory tracking performance was similar on both limbs in the HIV-only group, it was noticeably different for the HIV-stroke group, reflecting the presence of motor impairments in the non-dominant limb caused by stroke. The spatial span mean sequence length in each group was lower than the reported average span of 4.8 in a study that developed a computer-based version of the Corsi block-tapping task [42]. Given that moderate cognitive impairment may mask motor performance, the study subjects were further stratified by their cognitive and motor function.

**TABLE II.**
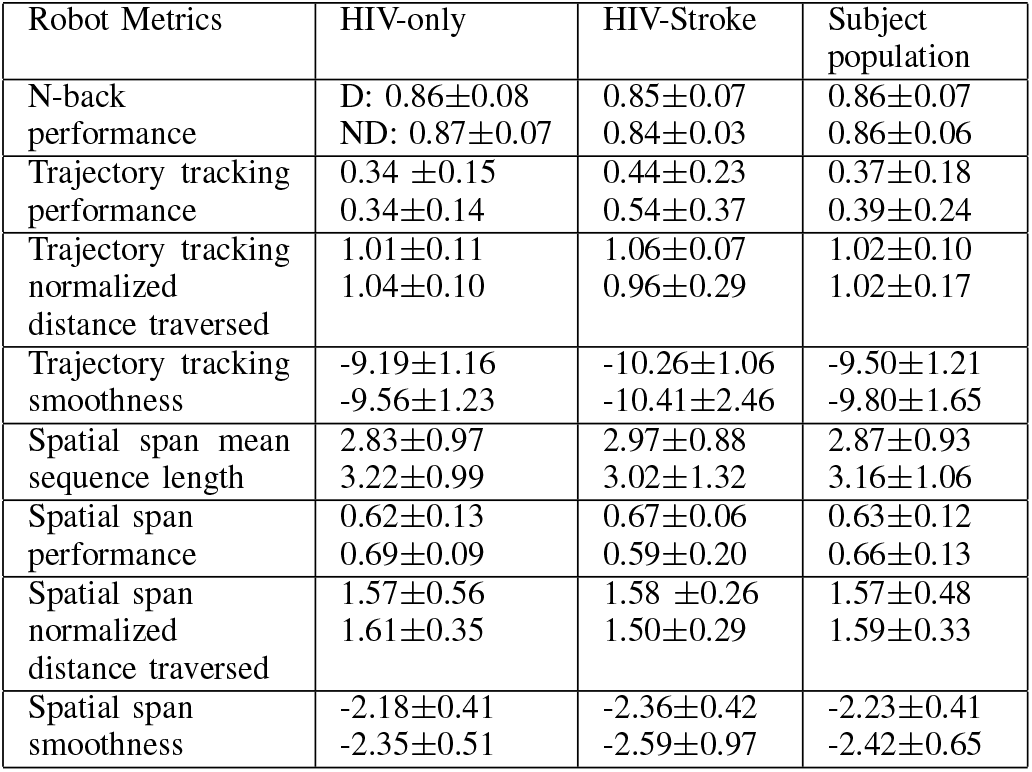
Group Robot Performance Results by Dominant (D) and Non-Dominant (ND) Limbs (Mean ± Standard Deviation)

| Robot Metrics | HIV-only | HIV-Stroke | Subject population |
| --- | --- | --- | --- |
| N-back performance | D: $0.86 \pm 0.08$<br>ND: $0.87 \pm 0.07$ | $0.85 \pm 0.07$<br>$0.84 \pm 0.03$ | $0.86 \pm 0.07$<br>$0.86 \pm 0.06$ |
| Trajectory tracking performance | $0.34 \pm 0.15$<br>$0.34 \pm 0.14$ | $0.44 \pm 0.23$<br>$0.54 \pm 0.37$ | $0.37 \pm 0.18$<br>$0.39 \pm 0.24$ |
| Trajectory tracking normalized distance traversed | $1.01 \pm 0.11$<br>$1.04 \pm 0.10$ | $1.06 \pm 0.07$<br>$0.96 \pm 0.29$ | $1.02 \pm 0.10$<br>$1.02 \pm 0.17$ |
| Trajectory tracking smoothness | $-9.19 \pm 1.16$<br>$-9.56 \pm 1.23$ | $-10.26 \pm 1.06$<br>$-10.41 \pm 2.46$ | $-9.50 \pm 1.21$<br>$-9.80 \pm 1.65$ |
| Spatial span mean sequence length | $2.83 \pm 0.97$<br>$3.22 \pm 0.99$ | $2.97 \pm 0.88$<br>$3.02 \pm 1.32$ | $2.87 \pm 0.93$<br>$3.16 \pm 1.06$ |
| Spatial span performance | $0.62 \pm 0.13$<br>$0.69 \pm 0.09$ | $0.67 \pm 0.06$<br>$0.59 \pm 0.20$ | $0.63 \pm 0.12$<br>$0.66 \pm 0.13$ |
| Spatial span normalized distance traversed | $1.57 \pm 0.56$<br>$1.61 \pm 0.35$ | $1.58 \pm 0.26$<br>$1.50 \pm 0.29$ | $1.57 \pm 0.48$<br>$1.59 \pm 0.33$ |
| Spatial span smoothness | $-2.18 \pm 0.41$<br>$-2.35 \pm 0.51$ | $-2.36 \pm 0.42$<br>$-2.59 \pm 0.97$ | $-2.23 \pm 0.41$<br>$-2.42 \pm 0.65$ |

### D. Stratification by Functional Subgroups (Hypothesis 1)

Figure 3 shows the distribution of the subject population by their functional groups using MoCA-EF subscores and BBT Z-scores to separate subjects by cognitive and motor function, respectively. The number of subjects in each of the four functional groups were the same when using dominant versus non-dominant BBT z-scores. There were two subjects in the low cognitive and low motor impairment group, five subjects in the low cognitive and moderate motor impairment group, six subjects in the moderate cognitive and low motor impairment group, and eight subjects in the moderate cognitive and moderate motor impairment group. Five HIV subjects and one stroke subject had different functional group classifications based on their dominant and non-dominant motor scores.

**Fig. 3.**
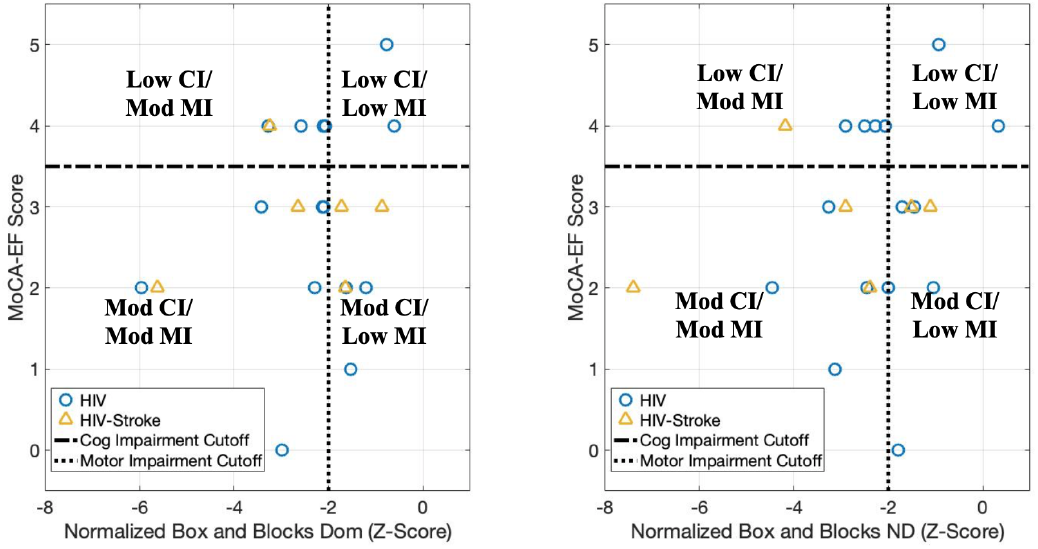
Distribution of subjects by cognitive and motor function, using a score of 3.5 for the MoCA EF cutoff and -2 as the BBT Z-score cutoff. The left figure is the distribution using the dominant limb BBT scores, while the right is from non-dominant limb BBT scores. (CI = cognitive impairment; MI = motor impairment; mod = moderate)

There was a statistically significant main effect of functional group on N-back performance (F(3,34) = 6.64, p = 0.001). There was no main effect of limb side or interaction effect. Subjects with low cognitive and low motor impairments performed better on the N-back task compared to subjects with moderate cognitive and moderate motor impairments (0.96 ± 0.01 vs. 0.83± 0.05, p = 0.001) and subjects with moderate cognitive and low motor impairments (0.96± 0.01 vs. 0.85± 0.06, p = 0.01). Fig. 4 (top) shows the N-back performance scores for each of the functional subgroups.

There was a statistically significant main effect of functional group on trajectory tracking performance error (F(3,34) = 7.78, p = 0.0004). There was no main effect of limb side or interaction effect. Subjects with moderate cognitive and moderate motor impairment had significantly higher performance error compared to subjects with low cognitive and moderate motor impairment (0.50± 0.25 vs. 0.28± 0.08, p = 0.04) and subjects with low cognitive and low motor impairment (0.50± 0.25 vs. 0.20± 0.04, p = 0.04). Fig. 4 (middle) shows the trajectory tracking performance for each of the functional groups.

**Fig. 4.**
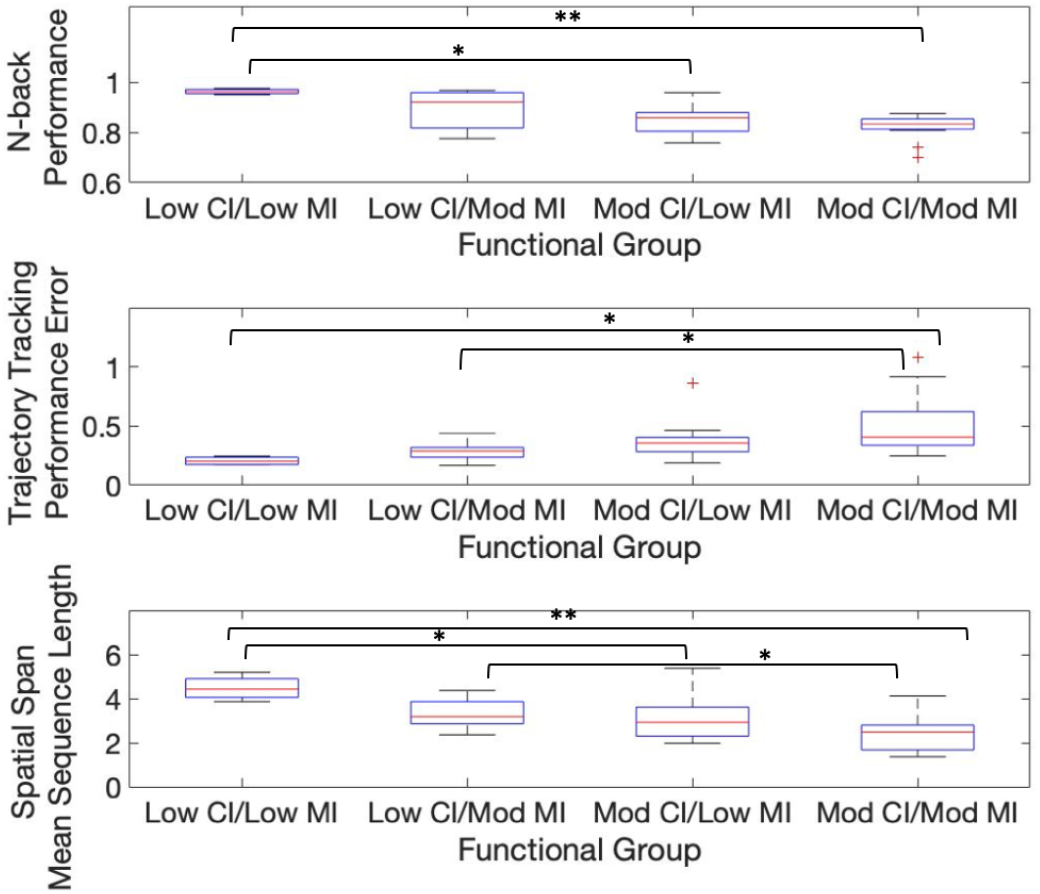
Box plots for each of the functional subgroups on N-back performance (top), trajectory tracking performance error (middle), and spatial span mean sequence length (bottom). CI = cognitive impairment; MI = motor impairment (* = p < 0.05, * * = p < 0.005 after correcting for multiple comparisons)

There was a statistically significant main effect of functional group on spatial span mean sequence length (F(3, 34) = 8.23, p = 0.0004). There was no main effect of limb side or interaction effect. Subjects with low cognitive and low motor impairment had longer average sequence lengths compared to subjects with moderate cognitive and moderate motor impairments (4.48± 0.56 vs. 2.39± 0.74, p = 0.0004) and subjects with moderate cognitive and low motor impairments (4.48± 0.56 vs. 3.12± 1.00, p = 0.04). Subjects with low cognitive and moderate motor impairment also had longer average sequence lengths compared to subjects with moderate cognitive and moderate motor impairment (3.31± 0.70 vs. 2.39± 0.74, p = 0.04). Fig. 4 (bottom) shows the spatial span mean sequence length for each of the functional subgroups.

There was a statistically significant main effect of functional group on spatial span performance, but no significant differences between any of the functional subgroups after correcting for multiple comparisons.

There were no statistically significant main or interaction effects for trajectory tracking normalized distance, trajectory tracking smoothness, spatial span normalized distance, or spatial span smoothness scores.

### E. Estimating clinical cognitive scores (Hypothesis 2)

#### 1) Dominant limb results

Fig. 5 shows the multiple linear regression models for each of the clinical cognitive assessments using dominant limb robot-based metrics as the predictors.

**Fig. 5.**
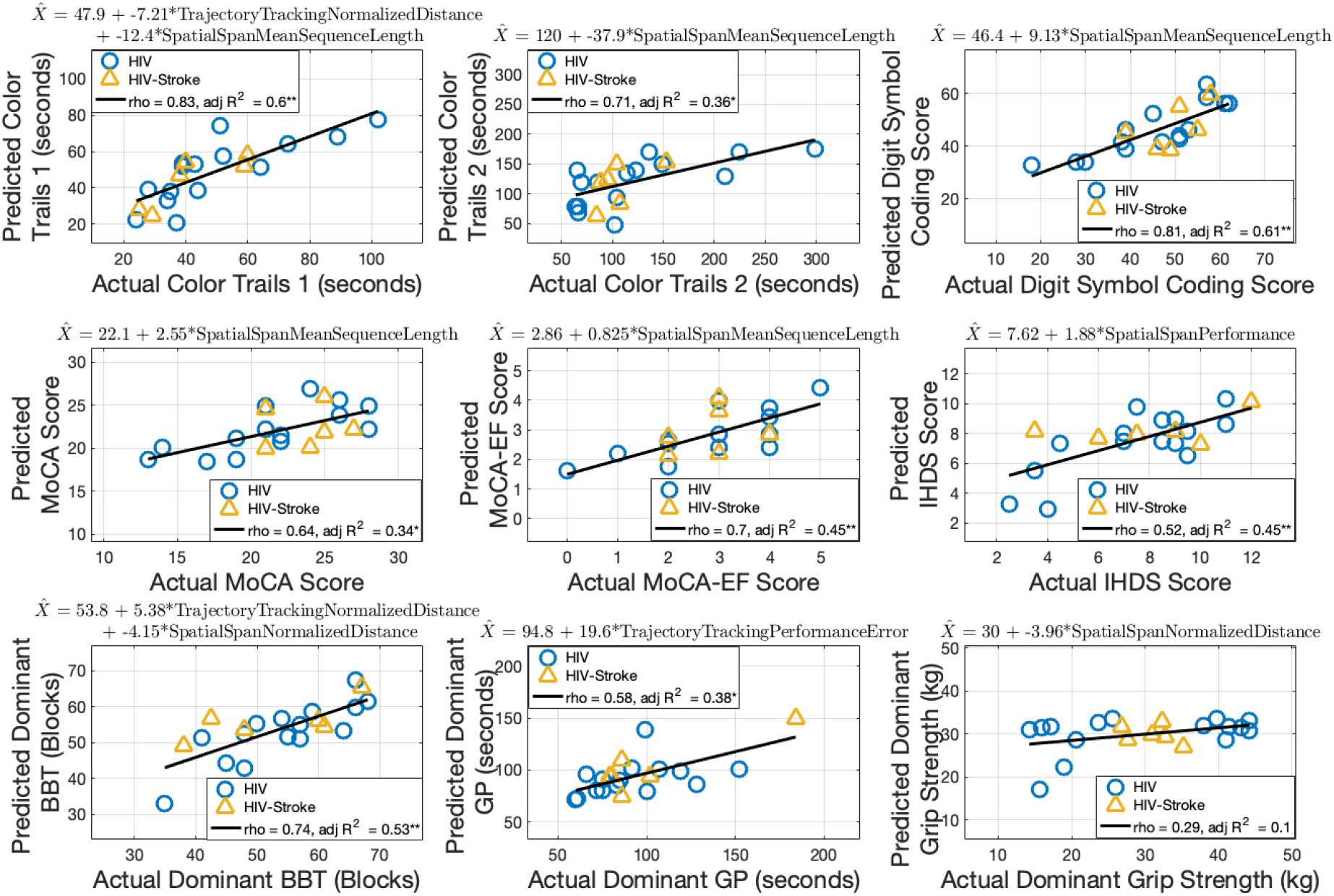
Multiple linear regression for clinical assessments using dominant limb robot-based metrics. The robot-based predictors for each model are included in the equation at the top of each subplot. Spearman’s rho and adjusted R^***2***^ are shown. (* **=** p < 0.05, * * **=** p < 0.001)

Color Trails 1 was predicted by a combination of trajectory tracking normalized distance traversed and spatial span mean sequence length (p = 0.03 and 0.001, respectively). The robot-based predictors accounted for 60% of the variance in the model, and the predicted scores strongly correlated with actual Color Trails 1 scores (rho = 0.83, p = 3.33× 10 ^*−*6^ ; adjusted R^2^ = 0.60, p = 1.13 ×10^*−*4^).

Color Trails 2 was predicted by spatial span mean sequence length (p = 0.002). The robot-based predictor accounted for 36% of the variance in the model, and the predicted scores strongly correlated with actual Color Trails 2 scores (rho = 0.71, p = 3.34×10^*−*4^; adjusted R^2^ = 0.36, p = 0.002).

Digit Symbol Coding was predicted by spatial span mean sequence length (p = 1.83 × 10^*−*5^). The robot-based predictor accounted for 61% of the variance in the model, and the predicted scores strongly correlated with actual Digit Symbol Coding scores (rho = 0.81, p = 7.06 × 10^*−*6^; adjusted R^2^ = 0.61, p = 1.83×10^*−*5^).

MoCA was predicted by spatial span mean sequence length (p = 0.003). The robot-based predictor accounted for 34% of the variance in the model, and the predicted scores moderately correlated with actual MoCA scores (rho = 0.64, p = 0.002; adjusted R^2^ = 0.34, p =0.003).

MoCA-EF was predicted by spatial span mean sequence length (p = 5.30 ×10^*−*4^). The robot-based predictor accounted for 45% of the variance in the model, and the predicted scores strongly correlated with actual MoCA-EF scores (rho = 0.70, p = 4.07× 10^*−*4^; adjusted R^2^ = 0.45, p = 5.30× 10^*−*4^).

IHDS was predicted by spatial span performance (p = 5.31× 10^*−*4^). The robot-based predictor accounted for 45% of the variance in the model, and the predicted scores moderately correlated with actual IHDS scores (rho = 0.52, p = 0.02; adjusted R^2^ = 0.45, p = 5.30× 10^*−*4^).

#### 2) Non-dominant limb results

Fig. 6 shows the linear regression models for each of the clinical cognitive assessments using non-dominant limb robot-based metrics as the predictors. Color Trails 1 was predicted by spatial span mean sequence length (p = 0.001). The robot-based predictor accounted for 39% of the variance in the model, and the predicted scores strongly correlated with actual Color Trails 1 scores (rho = 0.70, p = 3.73× 10^*−*4^; adjusted R^2^ = 0.39, p = 0.001).

**Fig. 6.**
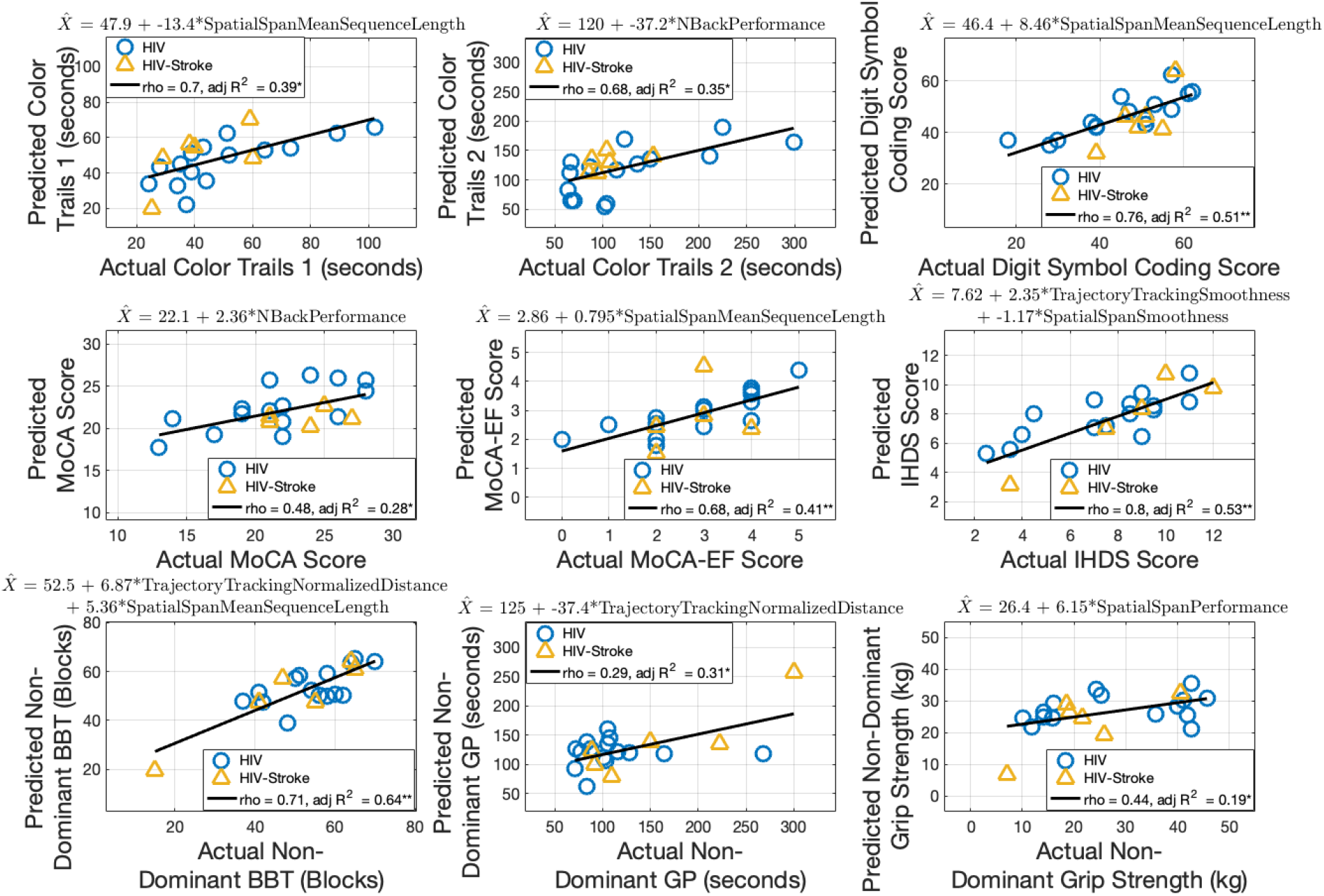
Multiple linear regression for clinical assessments using non-dominant limb robot-based metrics. The robot-based predictors for each model are included in the equation at the top of each subplot. Spearman’s rho and adjusted R^***2***^ are shown. (* **=** p < 0.05, * * **=** p < 0.001)

Color Trails 2 was predicted by N-back performance (p = 0.003). The robot-based predictor accounted for 35% of the variance in the model, and the predicted scores moderately correlated with actual Color Trails 2 scores (rho = 0.68, p = 7.79×10^*−*4^; adjusted R^2^ = 0.35, p = 0.003).

Digit Symbol Coding was predicted by spatial span mean sequence length (p = 1.51 × 10^*−*4^). The robot-based predictor accounted for 51% of the variance in the model, and the predicted scores strongly correlated with actual Digit Symbol Coding scores (rho = 0.76, p = 6.38× 10^*−*5^; adjusted R^2^ = 0.51, p = 1.51 ×10^*−*4^).

MoCA was predicted by N-back performance (p = 0.007). The robot-based predictor accounted for 28% of the variance in the model, and the predicted scores weakly correlated with actual MoCA scores (rho = 0.48, p = 4.07×10^*−*4^; adjusted R^2^ = 0.28, p = 0.007).

MoCA-EF was predicted by spatial span mean sequence length (p = 0.001). The robot-based predictor accounted for 41% of the variance in the model, and the predicted scores moderately correlated with actual MoCA-EF scores (rho = 0.68, p = 7.00×10^*−*4^; adjusted R^2^ = 0.41, p = 0.001).

IHDS was predicted by a combination of trajectory tracking smoothness and spatial span smoothness (p = 9.76× 10^*−*5^ and 0.02, respectively). The robot-based predictors accounted for 53% of the variance in the model, and the predicted scores strongly correlated with actual IHDS scores (rho = 0.80, p = 1.46×10^*−*5^; adjusted R^2^ = 0.53, p = 4.12×10^*−*4^).

### F Estimating clinical motor scores (Hypothesis 3)

#### 1) Dominant limb

Fig. 5 shows the linear regression models for each of the clinical motor assessments using dominant limb robot-based metrics as the predictors.

Dominant limb BBT was predicted bya combination of trajectory tracking normalized distance traversed and spatial span normalized distance traversed (p = 0.003 and 0.02, respectively). The robot-based predictors accounted for 53% of the variance in the model, and the predicted scores strongly correlated with actual BBT scores (rho = 0.74, p = 1.46 ×10^*−*4^; adjusted R^2^ = 0.53, p = 4.72 ×10^*−*4^).

Dominant limb GP was predicted by trajectory tracking performance (p = 0.002). The robot-based predictor accounted for 38% of the variance in the model, and the predicted scores moderately correlated with actual GP scores (rho = 0.58, p = 0.006; adjusted R^2^ = 0.38, p = 0.002).

Dominant limb grip strength was predicted by spatial span normalized distance traversed, but it was neither a significant predictor nor correlated to actual grip strength scores (rho = 0.29, p = 0.20; adjusted R^2^ = 0.10, p = 0.09).

### 2) Non-dominant limb

Fig. 6 shows the linear regression models for each of the clinical motor assessments using non-dominant limb robot-based metrics as the predictors.

Non-dominant limb BBT was predicted by a combination of trajectory tracking normalized distance traversed and spatial span mean sequence length (p = 0.002 and 0.01, respectively). The robot-based predictors accounted for 64% of the variance in the model, and the predicted scores strongly correlated with actual BBT scores (rho = 0.71, p = 3.41× 10^*−*4^; adjusted R^2^ = 0.64, p = 4.44 ×10^*−*5^).

Non-dominant limb GP was predicted by trajectory tracking normalized distance traversed (p =0.005). The robot-based predictor accounted for 31% of the variance in the model while the predicted scores were not significantly correlated with actual GP scores (rho = 0.29, p = 0.21; adjusted R^2^ = 0.31, p = 0.005).

Non-dominant limb grip strength was predicted by spatial span performance (p = 0.03). The robot-based predictor accounted for 19% of the variance in the model, and the predicted scores weakly correlated with actual grip strength scores (rho = 0.44, p = 0.04; adjusted R^2^ = 0.19, p = 0.03).

## IV. Discussion

### A. Gross motor impairments are prevalent in PLWHIV

This study aimed to use a robot-based approach to explore objective measures of cognitive and motor impairment in HIV and HIV-stroke populations. The HIV and HIV-stroke groups displayed no significant differences in clinical or robot-based scores. Subjects in both the HIV and HIV-stroke groups demonstrated mild to moderate impairment in executive function, information processing, and upper limb fine and gross motor domains relative to published population normal performance values in uninfected populations. These results are consistent with previous research demonstrating impairments in these domains in PLWHIV [6], [10], [43].

We found it notable that the HIV-only group demonstrated not only fine motor impairment as previously reported in the literature [7]–[9], [11], but also gross upper limb motor impairment. Gross motor impairment has generally been considered a pre-ART era manifestation of HIV infection, and studies since then have focused on the fine motor deficits that result from HIV [7]. Moderate bilateral gross motor impairment, as measured by the BBT and adjusted to healthy population norms, was present in 7 of 15 subjects in the HIV group. The prevalence of moderate bilateral fine motor impairment in the HIV-only subjects in this study (5 out of 15), as measured by the GP, is higher than what was reported in Wilson et al. (2 out of 12) in a group of PLWHIV with a similar average age of 57.9 years old [7]. These results suggest that gross upper limb motor impairments may be an overlooked effect of HIV and that the BBT can be used to identify these impairments as an alternative to the GP. This approach could be useful when examining patients with both HIV and stroke in particular, when motor impairments may be more prevalent [44].

### B. Robot-based metrics capture differences in functional subgroups

A wide range of impairments was observed in the subject population and there was no clear separation between the HIV and HIV-stroke groups on either the clinical assessments or robot-based metrics. As such, subjects were re-classified into one of four functional groups by their cognitive and motor performance. The results provide evidence in support of the study’s first hypothesis that robot-based metrics can differentiate subjects with and without moderate executive function or upper-limb motor impairments.

Subjects with moderate executive function impairment, regardless of motor status, performed worse on the N-back compared to subjects with low cognitive and low motor impairment. These results suggest the robot-based N-back can be used to isolate executive function deficits. This is consistent with previous findings that the paper-based N-back test, although specifically a test for working memory, engages executive function domains impacted by HIV [45].

Subjects with moderate executive function and moderate gross motor impairments performed worse on the robot-based trajectory tracking task compared to subjects with low cognitive impairment, regardless of motor status. This suggests that there might be a significant cognitive component to the trajectory tracking task that exacerbates performance error in the presence of executive function impairments.

Similarly to the robot-based N-back, subjects with moderate executive function impairment, regardless of motor status, had shorter sequences on the robot-based spatial span task compared to subjects with low cognitive and low motor impairment. Additionally, subjects with low cognitive and moderate motor impairment performed better than subjects with moderate cognitive and moderate motor impairment. These results suggest that a robot-based spatial span task can be used to detect executive function impairment, even in the presence of moderate motor impairment.

Together, these robot-based metrics provide a set of measures that are able distinguish between certain functional groups. Going forward, these represent a potential set of objective metrics that can be used to track longitudinal performance that relate to functional status in PLWHIV, stroke, and other conditions presenting with both motor and cognitive impairments.

### C. Robot-based metrics relate to HIV-related clinical assessments

To our knowledge, we are the first group to explore objective robot-based measures of both motor and cognitive impairments in PLWHIV. This study is a first step in developing more targeted neurorehabilitation strategies for PLWHIV exhibiting both motor and cognitive decline. The results support the study’s second hypothesis and show that both individual and linear combinations of robot-based metrics can successfully estimate clinical cognitive scores. The regression models for Color Trails 1, Digit Symbol–Coding, MoCA–EF and IHDS (adjusted R^2^ = 0.41–0.60) — excluding the non-dominant limb model for Color Trails 1 – performed the best, exceeding the effect size for which the study was powered. The robot-based measures also demonstrated statistically significant relationships with Color Trails 2 and MoCA.

This is one of the first studies to establish objective robot-based measures that relate to Digit Symbol–Coding, MoCA-EF subscores, or IHDS. Given that the Digit Symbol–Coding, MoCA-EF, and IHDS look at more specific cognitive domains related to executive function, this suggests the potential of robot-based metrics to identify more specific cognitive impairments going forward that are relevant to PLWHIV. Notably, the robot-based metrics that predicted these clinical scores were consistent with the robot-based metrics that showed differences between functional groups.

Two other studies that examine the relationship between robotic metrics and MoCA scores in stroke and traumatic brain injury populations reported correlation coefficients ranging between 0.49 and 0.65 that are similar to the values observed in this study (rho = 0.48–0.64) [21], [22].

The results provide evidence that robot-based metrics can successfully estimate clinical motor scores in PLWHIV. The dominant and non-dominant limb models for BBT scores (adjusted R^2^ = 0.53 and 0.64, respectively) performed the best, exceeding the effect size for which the study was powered and demonstrating strong correlations between predicted and actual scores. Using a multiple linear regression with eight robotic predictors derived from three tasks, Bosecker et al. demonstrated correlation coefficients between estimated and actual scores for the Fugl-Meyer, Motor Status Score, Motor Power Scale, and Modified Ashworth Scale of 0.42–0.80 on training models [23]. While the clinical motor metrics differed from those used in this study, these values were similar for the dominant and non-dominant BBT and GP models (rho = 0.31–0.74, respectively) with fewer predictors.

While computerized versions of the spatial span exist [42], the robotic aspect implemented in this study allows for kinematic measures to be observed that are reflective of motor function. This enables more detailed study of the interactions between cognitive and motor domains. The utility of this task can be seen by the high prevalence of metrics from this task demonstrating strong relationships with both cognitive and motor clinical scores.

### D. Relevance to HAND assessment, neurorehabilitation, global health, and robotics

Taken together, these results show the potential clinical utility of a robotics-based approach to assess motor and cognitive function in PLWHIV. Due to the involved nature of performing a complete HAND assessment, other alternatives have been explored to capture HIV-related neurocognitive impairments. For example, Fogel et al. used a stepwise multiple linear regression approach to predict a global deficit score (GDS) from a set of 24 metrics extracted from basic medical history in an older HIV population with an average age of 61.1±4.6 years, which was similar to the average of the HIV-only population in this study (56.2±5.4 years old) [46]. The GDS was calculated from a set of neuropsychological tests encompassing working memory and memory, motor, information processing, and learning domains that overlapped with some of the assessments in this study – specifically the GP, Trail Making A (equivalent to the Color Trails 1), and Digit Symbol–Coding. The ultimate three-term model from the Fogel et al. study had a R^2^ of 0.29, which is weaker compared to the R^2^ values for the Color Trails 1,Digit Symbol–Coding, and GP models in this study (R^2^ = 0.31–0.60) [46]. In Botswana, a lower-resource setting, a six-part neurocognitive battery, which also utilizes many of the same assessments as this study, was used to identify impairments in cognitive-motor areas in PLWHIV [8].

From a clinical rehabilitation perspective, increasing access to effective rehabilitation interventions and enhancing outcome measurement have been identified as research priorities in HIV, disability, and rehabilitation [47]. There is a need to develop interventions addressing the rapid aging and frailty associated with HIV to reduce disparities in health outcomes that can compound in the presence of other comorbidities or complications. No gold standard exists to capture the relationship between cognitive impairment and physical frailty as it relates to HIV [48]. While a limited number of studies have shown that physical exercise can induce improvements in physical, cognitive, and emotional wellbeing in both HIV and non-HIV populations, there is a need for further work to understand what impact exercise – including robot-based exercise – might have on the aging immune system in PLWHIV. A benefit to the objective quantification used in this study is the ability to track changes during the course of rehabilitation in specific metrics.

From a global health perspective, this technology-based approach provides a possible scalable strategy that is sensitive to subtle signs of functional decline. With more affordable rehabilitation robot systems becoming increasingly available, this approach has the potential to meet a huge rehabilitation need, particularly in lower resource settings where the capacity to supply additional rehabilitation professionals is lacking but the prevalence of non-communicable diseases necessitating rehabilitation is increasing [25]. This approach would be valuable particularly when medical history may be lacking or harder to assess. This preliminary work lays the groundwork for identifying specific impairments and developing HIV-specific neurorehabilitation strategies to address the various cognitive and motor impairments associated with aging with HIV. Our group is currently exploring this in Botswana.

From a robotics perspective, this study expands the application of rehabilitation robotics beyond stroke to PLWHIV and those living with cognitive impairments. Given that neurocognitive impairment is associated with instrumental ADL function [49], assessments and treatments should reflect the integration of both motor and cognitive domains that are often assessed in isolation. Like other robotic studies, large effect sizes were observed in this study, which can significantly reduce the sample size needed for clinical trials going forward [24]. This study also shows that clinical measures can be estimated from both limbs, which can be helpful in avoiding confounding factors, such as the presence of unilateral motor impairment that could result from stroke. This could be important to tune future neurorehabilitation strategies based on cognitive performance.

### E. Study limitations

A limitation of this study is the small sample size, which limits observations to moderate and strong relationships. While we observed strong correlations between robot-based measures and clinical cognitive and motor assessments relevant to the HIV population, correlation studies are susceptible to the distribution of the data across span of the predictor variables. While we had adequate distribution across many variables, we were not able to get an even distribution across functional groups, which could have biased the analysis. Despite these limitations, further studies with a larger sample size and a longitudinal evaluation of this approach is warranted.

## Data Availability

De-identified data is available upon request, and will be available as a repository soon.

## V. Acknowledgements

This work was made possible through core services and support from the National Institute Of Neurological Disorders and Stroke of the National Institutes of Health (T32NS091006); the University of Pennsylvania’s Center for AIDS Research (P30AI 045008); the University of Pennsylvania’s Center for Biomedical Image Computing and Analytics; and the University of Pennsylvania’s Departments of Bioengineering and Physical Medicine and Rehabilitation. The content is solely the responsibility of the authors and does not necessarily represent the official views of the National Institutes of Health. The authors would like to acknowledge Kristine Lima and Christine Kurian for their contributions to the study.

## Notes

### Competing Interest Statement

Michelle Johnson is a co-founder of Recupero Robotics, LLC, which is commercializing the rehabilitation robot system used in the study.

### Clinical Trial

Study was not a clinical trial

### Author Declarations

University of Pennsylvania IRB Protocol #823511

